# Misleading and avoidable: design-induced biases in observational studies evaluating cancer screening—the example of site-specific effectiveness of screening colonoscopy

**DOI:** 10.1101/2024.04.29.24306522

**Authors:** Malte Braitmaier, Sarina Schwarz, Vanessa Didelez, Ulrike Haug

## Abstract

Observational studies evaluating the effectiveness of cancer screening are often biased due to non-alignment at time zero, which can be avoided by target trial emulation (TTE). We aimed to illustrate this by evaluating site-specific effectiveness of screening colonoscopy regarding colorectal cancer (CRC) incidence.

Based on a German health care database, we assessed the effect of screening colonoscopy vs. no screening colonoscopy in preventing CRC in the distal and the proximal colon over 12 years of follow-up in 55–69-year-old persons. We compared four different study designs: cohort and case-control study, each with/without alignment at time zero.

In both analyses with time zero-alignment, screening colonoscopy showed a rather similar effectiveness in reducing the incidence of distal and proximal CRC (cohort analysis: 32% (95% CI: 27-37%) vs. 28% (20-35%); case-control analysis: 27% vs. 33%). Both analyses without alignment suggested a difference by site: Incidence reduction regarding distal and proximal CRC, respectively, was 65% (61-68%) vs. 37% (31-43%) in the cohort analysis and 77% (67-84%) vs. 46% (25-61%) in the case-control analysis.

Violations of basic design principles can substantially bias the results of observational studies. In our example, it falsely suggested a much stronger preventive effect of colonoscopy in the distal vs. the proximal colon. Our study illustrates that TTE avoids such design-induced biases.

## Introduction

Randomized controlled trials (RCT) are the gold standard for evaluating the effectiveness of cancer screening. However, existing RCTs in this field do not answer all relevant research questions. For screening colonoscopy, for example, an RCT has recently been published (NordICC trial) demonstrating its effectiveness in reducing colorectal cancer (CRC) incidence overall^1^, but it was not powered to compare the effectiveness in the distal vs. the proximal colon.

Complementary evidence from observational studies is therefore needed. Apart from potential confounding, there is a high risk of bias and thus of misleading results if such studies are inadequately designed. Indeed, several observational studies have reported a markedly stronger preventive effect of screening colonoscopy in the distal as compared to the proximal colon^2–4^, while a cohort study designed following the principle of target trial emulation (TTE) showed a similar effectiveness of screening colonoscopy in the distal and the proximal colon^5^. We argued that the difference by site in the former studies was due to biases induced by non-alignment at “time zero”, i.e. at baseline. This means that I) the assessment of eligibility, II) the assignment to study arms and III) the start of follow-up were not aligned as they would be in an RCT and as it would be ensured in an observational study designed based on the principle of TTE^6^. Specifically, previous studies often defined exposure based on pre- or post-baseline information on colonoscopy. As we further argued, this lack of alignment in previous studies led to overestimating the effectiveness of screening colonoscopy. Due to the different age pattern of distal and proximal CRC, this bias affected distal CRC more than proximal CRC, i.e. the difference in effectiveness by site was an artefact.

To demonstrate this, we compared different study designs with and without alignment at time zero aiming to investigate the question of site-specific effectiveness of screening colonoscopy in reducing CRC incidence. For the two designs without alignment we used a cohort study design, where the assignment to study arms occurs *before* time zero (pre-baseline), and a nested case control study design, where the assignment to study arms occurs *after* time zero (post-baseline). For the two designs with alignment, assignment to study arms was done based on screening colonoscopy at baseline. The current paper is part of a growing literature identifying violations of alignment at time zero as a potential source of major bias in observational studies^6–8^.

## Methods

### Data source and study population

We used the German Pharmacoepidemiological Research Database (GePaRD) which comprises claims data from four statutory health insurance providers in Germany and covers about 20% of the German population^9^. In GePaRD, information on utilization of screening colonoscopy, offered in Germany to persons aged 55 or older since October 2002 (since April 2019 also offered to men aged 50-54) with a screening interval of 10 years, is distinguishable from diagnostic colonoscopy. Each year, 2-3% of the target population utilize screening colonoscopy, yielding a cumulative participation of 20-30%^10^. As previously described, the data source enables the valid identification of incident CRCs^11^. Furthermore, it contains appropriate information to apply in- and exclusion criteria and to adjust for confounding as relevant to the research question on the effectiveness of (at least one) screening colonoscopy in reducing CRC incidence^5^. For the present study, we used data from 2004 to 2020.

Based on this data source, we applied four different study designs to address the research question, specifically a cohort and a case-control study design, each with and without alignment at time zero. The study designs without alignment at time zero were inspired by published examples^2,12–14^, and were partly complemented by sensitivity analyses. For each of these four studies, persons were selected from the same population. Specifically, the source population was a cohort of persons aged 55–69 at baseline, who were continuously insured for at least three years before baseline.

### Cohort study without alignment at time zero

The cohort started in 2009 (baseline). Similar to a previous study^2^, individuals were assigned to the screening colonoscopy arm if they had a screening colonoscopy any time before baseline, including the baseline quarter. Individuals were assigned to the control arm if they did not undergo screening colonoscopy any time before baseline, including the baseline quarter. In a sensitivity analysis, we considered both screening and diagnostic colonoscopies for the assignment to the study arms, because some of the previous studies did not distinguish between these examinations. Eligibility criteria were checked at baseline (among others, prevalent CRC as exclusion criterion) and the outcome variable (incident CRC) was assessed beginning with baseline (start of follow-up). Persons were followed up until end of study period (end of 2020), end of continuous insurance coverage, death or CRC diagnosis, whichever occurred first. We also conducted sensitivity analyses starting the cohort in 2010 and 2011, respectively.

When using such a study design, the assessment of eligibility and the start of follow-up are aligned, but the assignment to the screening and the control arm is based on a period *before* time zero (pre-baseline). Specifically, individuals in the colonoscopy arm had the examination in the past (i.e. they were assigned to the screening arm based on past exposure) rather than *at* time zero.

### Cohort study with alignment at time zero

As described previously^5^, we emulated sequential trials for each calendar quarter from 2007 to 2011. The emulation of sequential target trials makes full use of the information from longitudinal data without violating principles of study design by using pre- or post-baseline information for the assignment to study arms. At the baseline quarter of each trial, eligibility was assessed (among others, individuals with prevalent CRC or previous colonoscopy were excluded). Eligible individuals were then assigned to the screening arm if they underwent a screening colonoscopy *in the baseline quarter* of the respective trial (i.e. irrespective of participation in a second screening colonoscopy offered 10 years later) and to the control arm otherwise. Individuals were followed up until end of study period (end of 2020, i.e. follow-up was longer than in our previous analysis), end of continuous insurance coverage, death or CRC diagnosis, whichever occurred first. This study design made sure that assessment of eligibility criteria, assignment to the screening and control arm, and start of follow-up were aligned at time zero as would be the case in an RCT.

### Case-control study without alignment at time zero

We applied a case-control design frequently used in the published literature^12–17^. Essentially, CRC cases are identified (date of diagnosis corresponds to index date) and matched with controls free of CRC at index date. Then screening colonoscopy use *ever before* or within a certain time period *before* the index date is assessed in cases and controls, i.e. colonoscopies leading to CRC diagnosis are not considered as exposure in this type of study. Here, we selected all individuals from the source population entering the cohort in 2009 with a CRC diagnosis in 2018-2020. For each case we matched up to five controls on age (+/- one year) and sex (sampling without replacement). The exposure variable was then defined as any screening colonoscopy between 2009 and the index date, i.e. exposure to colonoscopy use was assessed within 10-12 years *before* the index date. Colonoscopies conducted in the six months before CRC diagnosis were not considered in defining the exposure. As mentioned above, this approach corresponds to published case-control studies which ignore colonoscopies conducted as part of the diagnostic process leading to the current diagnosis^12–17^. In general, it is a fundamental characteristic of traditional case-control studies to assess exposure before disease onset. In a sensitivity analysis, we considered both screening and diagnostic colonoscopies for the assignment to exposure groups. Again, we also conducted sensitivity analyses using the years 2010 and 2011 for cohort entry, i.e. the source population underlying this nested case-control study.

In the case-control design we used here (nested within a cohort with a clear baseline), the assessment of eligibility and the start of follow-up were aligned. The assignment to the screening and the control arm depended on whether or not screening colonoscopy was utilized during follow-up, and thus occurred *after* time zero (post-baseline) instead of *at* time zero. Note that in case-control studies not nested in a cohort, there typically are additional misalignments ^12,15^. Specifically, eligibility is assessed at index date. Furthermore, the start of follow-up (baseline) is unclear, i.e. there is no alignment and the distinction between pre- and post-baseline information is void.

### Case-control study with alignment at time zero

Following the approach described by Dickerman et al. ^18^, a case-control study was nested within the original cohort of sequential emulated target trials, and colonoscopy use was assessed at baseline of each emulated trial. We included CRC patients with an incident CRC diagnosis at any point during follow-up (until 2020) and then used risk set sampling to match up to five controls to each case. We sampled matched controls with replacement, i.e. the same control could be matched to more than one case. Matching variables were the same as above. The key difference to the case-control study without alignment is that exposure assignment was based on information available at the start of the emulated trial, i.e. at time zero, instead of information occurring after time zero. This approach has been shown to avoid self-inflicted biases in the same way as a prospective study using TTE^18^.

### Data analysis

Time was discretized into units of calendar quarters (i.e. three-month time intervals), since some information in the database (outpatient diagnosis codes) is only available on a quarterly basis. We estimated the effect on site-specific CRC incidence, i.e. the outcome was the first CRC diagnosis during follow-up. A CRC diagnosis at the other site or with unknown location and death from any cause were regarded as competing events. We estimated the site-specific total effects, i.e. without elimination of competing events (see Young et al. ^19^ for details on total and controlled direct effects in competing risk survival settings). As shown in our initial study, treating death as a censoring event did not substantially change the effect estimates given the included age range (55-69 years). ^5^ For the cohort studies, we estimated site-specific cumulative incidence functions (CIF) via pooled logistic regressions, which were adjusted for baseline confounders via inverse probability of treatment weighting (see Supplement 1 for details on the pooled logistic regression). Adjustment variables were: Age, sex, educational attainment, type 2 diabetes, codes indicative of alcohol abuse, codes indicative of nicotine dependence, antiplatelet therapy, use of preventive services, menopausal hormone therapy, coded family history of CRC. Effects were estimated as adjusted relative risks (RR) at the end of follow-up based on these CIFs. As previously shown, adjustment yielded satisfactory covariate balance and a negative control analysis did not indicate any residual confounding^5^. For the cohort study with alignment at time zero, our effect estimate corresponds to the observational analog of the intention-to-treat effect. This avoided additional statistical assumptions, which we preferred given our methodological focus, while recognizing that a per-protocol effect would be more relevant in other research contexts. Controls (i.e. persons with no screening colonoscopy at baseline) may have undergone screening colonoscopy during follow-up but as reported in our previous study, this had no relevant impact on our results: The proportion of CRCs among persons in the screening arm who were also included as controls in at least one previous trial was low and similar in the distal and the proximal colon (4.2% and 4.8%)^5^. To further support this, we conducted additional analyses estimating the per-protocol effects for the cohort design with alignment, comparing screening at baseline with never screening colonoscopy (Supplement 2). Confidence intervals were estimated via person-level bootstrap. For the case-control studies, effects were estimated as adjusted odds ratios (ORs) obtained via conditional logistic regression. For the case-control analysis with alignment, no confidence intervals could be obtained due to computational limitations: The emulation of sequential trials with repeated cohort entry would require bootstrapping of the underlying study population, where matching is repeated for every bootstrap sample^18^, which resulted in unfeasible run times in this particular analysis with the IT equipment available to us. Bootstrapping the matched cohort would not yield valid confidence intervals^20^.

## Results

### Cohort study without alignment at time zero

We selected a random sample of 200,000 individuals in the control arm and 200,000 individuals in the screening colonoscopy arm. The adjusted relative risk after 12 years of follow-up was 0.35 (95% CI: 0.32-0.39) for distal CRC and 0.63 (95% CI: 0.57-0.69) for proximal CRC (Table 1). The adjusted cumulative incidence curves are given in Fig. **1**. As shown in Supplement 3, results were similar when the year 2010 or the year 2011 was used as baseline. In sensitivity analyses considering both screening and diagnostic colonoscopies as exposure, the adjusted 12-year relative risk was 0.40 (95% CI: 0.37-0.44) for distal CRC and 0.66 (95% CI: 0.60-0.72) for proximal CRC (Supplement 4).

**Fig. 1:**
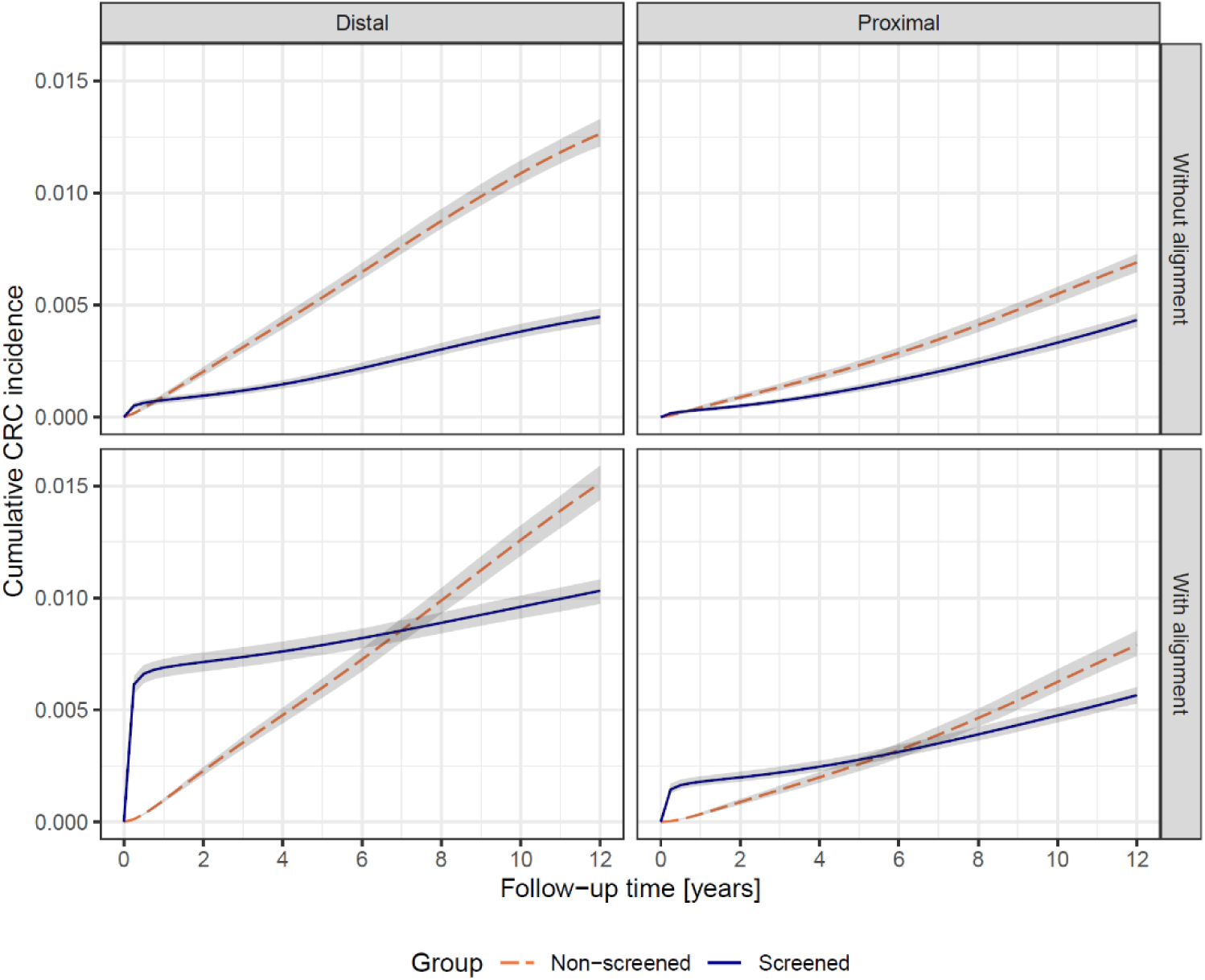
Adjusted cumulative incidence functions for distal (left column) and proximal (right column) CRC from the cohort study design with alignment at time zero (bottom row) and the cohort study design without alignment at time zero (top row)

**Table 1:** Results of cohort study designs without and with alignment at time zero (adjusted for baseline covariates).

|  | Control group | Screening group | Adjusted relative risk | (95% CI) |
| --- | --- | --- | --- | --- |
| **Design without alignment at time zero** |  |  |  |  |
| Number at risk | 200,000 | 200,000 |  |  |
| Number of CRC cases |  |  |  |  |
| Distal CRC | 2,472 | 829 | 0.35 | (0.32-0.39) |
| Proximal CRC | 1,290 | 823 | 0.63 | (0.57-0.69) |
| **Design with alignment at time zero** |  |  |  |  |
| Number at risk | 1,241,071 | 192,054 |  |  |
| Number of CRC cases |  |  |  |  |
| Distal CRC | 16,750 | 1,678 | 0.68 | (0.63-0.73) |
| Proximal CRC | 8,548 | 919 | 0.72 | (0.65-0.80) |

### Cohort study with alignment at time zero

Overall, 192,054 persons were included in the screening colonoscopy arm. The 5% random sample (restriction due to computational limitations) of controls assigned to the no screening arm included 116,452 persons (1,241,071 non-unique). The adjusted relative risk after 12 years of follow-up was 0.68 (95% CI: 0.63-0.73) for distal CRC and 0.72 (95% CI: 0.65-0.80) for proximal CRC (Table 1). Figure 1 shows the adjusted cumulative incidence curves for distal and proximal CRC. The distribution of screen-detected and post-colonoscopy CRCs (i.e. non-screen-detected CRCs) by site is shown in Supplement 5. As shown in Supplement 2, additional analyses estimating the per-protocol effect for the cohort design with alignment (comparison of screening at baseline vs. never screening colonoscopy) showed similar results.

### Case-control study without alignment at time zero

Overall, 446 cases with distal CRC matched to 2,230 controls and 302 cases with proximal CRC matched to 1,510 controls were included. The adjusted ORs for distal and proximal CRC were 0.23 (95% CI: 0.16-0.33) and 0.54 (95% CI: 0.39-0.75), respectively (Table 2). When the year 2010 or the year 2011 was used to define the source population, the difference by site was similar (Supplement 3). The sensitivity analysis considering both screening and diagnostic colonoscopy as exposure yielded similar results; the adjusted ORs for distal and proximal CRC were 0.20 (95% CI: 0.15-0.26) for distal CRC and 0.44 (95% CI: 0.33-0.58) for proximal CRC, respectively (Supplement 4).

**Table 2:** Results of case-control study designs without and with alignment at time zero.

| Site | Case status |  | Adjusted OR |  |
| --- | --- | --- | --- | --- |
|  | Cases | Controls | (95% CI)<sup>§</sup> |  |
| Design without alignment at time zero |  |  |  |  |
| Number of distal CRCs / controls | 446 | 2,230 |  |  |
| Thereof exposed | 36 | 653 | 0.23 | (0.16-0.33) |
| Number of proximal CRCs / controls | 302 | 1,510 |  |  |
| Thereof exposed | 54 | 434 | 0.54 | (0.39-0.75) |
| Design with alignment at time zero |  |  |  |  |
| Number of distal CRCs / controls | 19,081 | 93,650 |  |  |
| Thereof exposed | 1,708 | 12,803 | 0.70 |  |
| Number of proximal CRCs / controls | 9,916 | 49,065 |  |  |
| Thereof exposed | 969 | 6,669 | 0.72 |  |

<sup>§</sup>: Confidence intervals could not be obtained for the case-control analysis with alignment at time zero due to computational limitation (see methods section).
OR: Odds Ratio

### Case-control study with alignment at time zero

Overall, 19,081 cases with distal CRC matched to 93,650 controls and 9,916 cases with proximal CRC matched to 49,065 controls were included. The adjusted ORs for distal and proximal CRC were 0.70 and 0.72, respectively (Table 2).

A table of baseline covariates by study design is given in Supplement 6.

## Discussion

To the best of our knowledge, our study is the first to systematically compare different study designs to assess the effectiveness of screening colonoscopy in reducing CRC incidence in the distal vs. the proximal colon. Our cohort and case-control analyses with alignment at time zero showed no relevant difference in the effectiveness by site. Using study designs without alignment at time zero led to an overestimation of the effectiveness of screening colonoscopy overall. The overestimation affected distal CRCs considerably more than proximal CRCs, i.e. purely by design there appeared to be a difference in effectiveness by site. This finding held up in sensitivity analyses varying data years and the type of examinations considered for the exposure definition (only screening or also diagnostic colonoscopy). Our findings demonstrate that the difference in the effectiveness of colonoscopy by site reported by previous observational studies was due to bias introduced by inadequate study design.

As illustrated in Supplement 7 using directed acyclic graphs, the bias underlying studies using pre-baseline information on colonoscopy for the assignment to study arms can be expressed as a form of collider stratification bias^21,22^. To give an intuitive explanation, let us revisit the study by Guo et al. ^2,5^: At baseline, patients were asked about past colonoscopy use and— based on this information—assigned as exposed or unexposed to colonoscopy. Persons reporting a prior CRC diagnosis at baseline were excluded^2^. Given that colonoscopy is one of the main tools by which CRC is diagnosed, this process removes individuals with previously diagnosed CRC from the exposed group, i.e. it enriches the exposed group with individuals who are known to be free of CRC. No such selection process takes place in the unexposed group. This leads to a lower prevalence of preclinical CRC at baseline in the exposed as compared to the unexposed group. As a consequence, this selection reduces the number of CRCs occurring during follow-up in the exposed group as compared to the unexposed group and thus leads to overestimation of the effect of screening on CRC incidence. This is also evident from Figure 1, showing that the absolute risk in the screened group is much lower without alignment at time zero as compared to the design with alignment. As the vast majority of CRCs diagnosed at an age when persons are typically included into screening studies are in the distal colon^23^ while proximal CRCs become more common at older age, this bias mainly affects results for distal CRC, i.e. as mentioned above there appeared to be a difference in effectiveness by site purely by design. We note that in addition to the initial exposure assignment, Guo et al. also used an updated exposure variable in a Cox model with time-dependent covariates. However, this does not correct the initial selection issue at the start of follow-up. Further, including time-updated covariate information in the outcome model of a time-dependent exposure may introduce bias due to exposure-covariate feedback^24^. Moreover, hazard-based effect measures are discouraged in causal analyses due to the built-in selection bias ^19,25,26^.

The above argument applies to studies using pre-baseline information for the assignment to exposure groups. Many other studies used post-baseline information for the assignment to exposure groups, also inducing bias. We illustrated this by the case-control study without alignment at time zero: Whenever after baseline CRC is detected in a person at his or her first colonoscopy, as is the case for most screen-detected CRCs, this person is assigned to the unexposed group as there was no previous colonoscopy and the actual colonoscopy detecting the CRC is not considered as prior exposure. This enriches the unexposed group with CRCs and thus leads to overestimation of the effectiveness of screening. As the majority of screen-detected CRCs are in the distal colon, this bias predominantly affects CRCs in the distal colon and thus leads to an artificial difference in the effectiveness of colonoscopy by site (see also Supplement 8). In our case-control study design embedded in an emulated target trial with alignment at time zero, in which screen-detected CRCs are correctly assigned, no relevant difference in the effectiveness of colonoscopy by site was observed. Of note, misalignment due to post-baseline exposure assignment is typical of but not limited to case-control designs on cancer screening. It can also occur in inadequately designed cohort studies and is not overcome by using a time-varying exposure variable in a hazard model. This is explained in more detail in Supplement 9 based on the example of the study by Nishihara et al. ^3^

In summary and more generally, both study designs without alignment at time zero have in common that there are mechanisms that lead to inappropriate consideration of screen-detected CRCs, i.e. in the screening arm there was no peak in CRC incidence immediately after baseline as it would be the case in an RCT. Of course, this overestimates the impact of screening on CRC incidence, particularly for distal CRC, as illustrated in Figure 1. The flawed approaches ignore the fact that a screening colonoscopy sometimes comes too late to prevent CRC. Following the publication of the NordICC study, there was a discussion whether it is appropriate to include persons with preclinical CRC, causing the peak at baseline, in a prevention trial^27,28^. However, from a public health perspective, it is important to also take into account CRCs that are not prevented by screening in order to avoid overestimating the effectiveness of CRC screening at the population level. Apart from this, it should be noted that studies without alignment at time zero do not provide a valid answer to the question regarding the size of the preventive effect of colonoscopy in persons free of CRC at baseline.

It should be noted that, although we focus our discussion on biases most relevant for site-specific effectiveness of screening colonoscopy, misalignment at time zero should also be avoided for many other reasons. Our examples for using pre- and post-baseline information for the assignment to exposure groups were selected according to their relevance to our research question but there are certainly other types misalignment. Rasouli et al. ^29^ demonstrated that time related issues such as prevalent user bias or time-varying confounding are a threat to case-control designs not embedded in an emulated target trial. Also, Dickerman et al. showed—based on case-control studies investigating the impact of statins on CRC risk— the biases inherent to traditional case-control studies and the potential of avoiding bias and wrong conclusions if the study is designed following the principle of TTE^18^. Similarly, there are many examples of biases other than those we discussed here that are inherent to cohort studies without alignment at time zero^8^.

Our findings have several implications. First, regarding research on CRC screening, previous studies suggesting a lower effectiveness of colonoscopy in the proximal colon stimulated a search for reasons that may explain the occurrence of post-colonoscopy CRCs specifically in the proximal colon. It was suggested that one main reason relates to sessile serrated lesions as they are more difficult to detect and more often occur in the proximal colon^30^. While we do not question the important role of these lesions, our findings may encourage a broadening of the discussion of potential reasons leading to post-colonoscopy CRCs. Indeed, in our emulated target trial on screening colonoscopy, the proportion of post-colonoscopy CRCs located in the distal vs. the proximal colon was rather similar (Supplement 5). A one-sided focus on lesions that occur more frequently in the proximal colon therefore seems too narrow regarding the identification of lesions possibly leading to post-colonoscopy CRCs.

Our results also have implications beyond the specific research question of our study. Observational data are often used to evaluate the effectiveness of cancer screening. They represent a valuable data source to complement RCT evidence in this field, as RCTs on cancer screening are scarce, were often conducted many years ago and are typically not powered to estimate, for example, subgroup-specific effects or differences by cancer subtype. However, our study illustrates that—in addition to appropriate control of confounding—it is of key importance to design these studies in a way to ensure alignment at time zero. This means that assessment of eligibility, assignment to the screening and control arm and start of follow-up must be aligned. Otherwise, there is a high risk of bias. Weighing the advantages and disadvantages of the two study designs with alignment at time zero, we generally recommend a cohort design for several reasons, some of which are: 1) The computational cost of embedding a case-control study in an emulated target trial (cohort study) is higher than simply conducting the emulated target trial itself. 2) The matching of cases to controls leads to a loss of sample size among controls, possibly lowering statistical efficiency. However, a case-control design with alignment might be useful and cost-efficient if the available data source lacks pivotal information and needs to be substituted by additional data (e.g. biomarkers measured in baseline samples).

Specific strengths of our study include the systematic comparison of different study designs as well as the comprehensive sensitivity analyses. Given that all analyses were conducted using the same data source and referred to the same setting, there is no heterogeneity regarding, for example, the study variables or setting-related factors such as the uptake of surveillance colonoscopy or colonoscopy quality. This strengthens our conclusion that differing results of the analyses with and without alignment at time zero are exclusively due to the study design.

It should be noted that our findings apply to the population aged 55-69, covering the typical screening age range of CRC screening. Whether screening colonoscopy is equally effective in the distal and proximal colon in older age groups cannot be answered by our study, nor did we address the endpoint CRC mortality. These research questions were beyond our study’s scope, as our primary objective was to illustrate the relevance of design-induced biases and the possibility to avoid them using TTE, exemplified by investigating site-specific effectiveness of screening colonoscopy in reducing CRC incidence.

In conclusion, our study demonstrates that violation of alignment at time zero can substantially bias the results of observational studies on cancer screening. In our example, it falsely suggested an almost doubled preventive effect of colonoscopy in the distal vs. the proximal colon. The difference disappeared when the same data were analyzed using a TTE approach, which is known to avoid design-induced biases.

## Data Availability

As we are not the owners of the data we are not legally entitled to grant access to the data of the German Pharmacoepidemiological Research Database. In accordance with German data protection regulations, access to the data is granted only to employees of the Leibniz Institute for Prevention Research and Epidemiology - BIPS on the BIPS premises and in the context of approved research projects. Third parties may only access the data in cooperation with BIPS and after signing an agreement for guest researchers at BIPS.

## Supplement 1: Pooled logistic regression

In our prospective study designs, we used pooled logistic regression to estimate the cumulative incidence of colorectal cancer (see Robins et al. for details on this approach ^1^). We chose this analytical approach because it allowed us to compare the risk of CRC throughout follow-up and avoided (unjustified) proportional hazards assumptions. First, we estimated inverse probability of treatment weights (IPTW) to adjust for baseline confounding. For this, we fitted a logistic model with the exposure group as dependent variable and baseline covariates as explanatory variables. This model yielded an estimated probability of being assigned to the exposed group 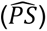 which was used to estimate IPTW weights as 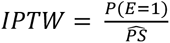 for the exposed group and 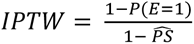 for the unexposed group, with 𝐸 ∈ {0,1} being the exposure groups.

Next, we fitted a pooled logistic regression model on the long-format data (i.e. one data row per (possibly non-unique) person per time point), which took the form

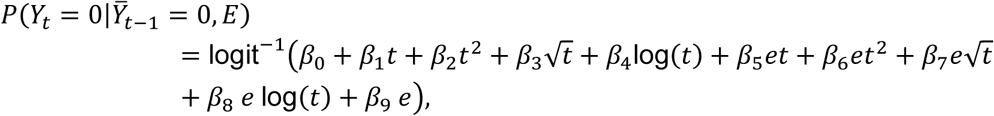

with 𝑡 being follow-up time and 𝑒 being the observed exposure value. The above model was fitted on the weighted data and predicted probabilities of no event occurring at time 𝑡 were extracted per exposure group. Cumulative incidence functions were then estimated as one minus the cumulative product of these predicted probabilities over time.

## Supplement 2: Cohort study with alignment – results of the per-protocol analysis

In the main manuscript, we reported results for the effect of screening colonoscopy at baseline versus no screening colonoscopy at baseline, i.e. without enforcing adherence to a sustained never screening strategy via artificial censoring. In this supplement, we report per-protocol results for the cohort design with alignment at time zero, comparing screening colonoscopy at baseline with never screening. Non-unique individuals were artificially censored in the control group, if and when they underwent screening colonoscopy during follow-up (this affected 19.8 %). Inverse probability of censoring weights were applied to adjust for resulting selection effects. We used the same (but time-updated) covariates that we used in the baseline propensity model to estimate the probability of being artificially censored.

The adjusted per-protocol cumulative incidence functions for incident distal and proximal CRC are given in Figure S5. The adjusted relative risks at 12 years were 0.74 for distal and 0.76 for proximal CRC. As with the main analysis, we did not observe notable differences in the relative effect of screening colonoscopy by CRC location.

**Figure S5:**
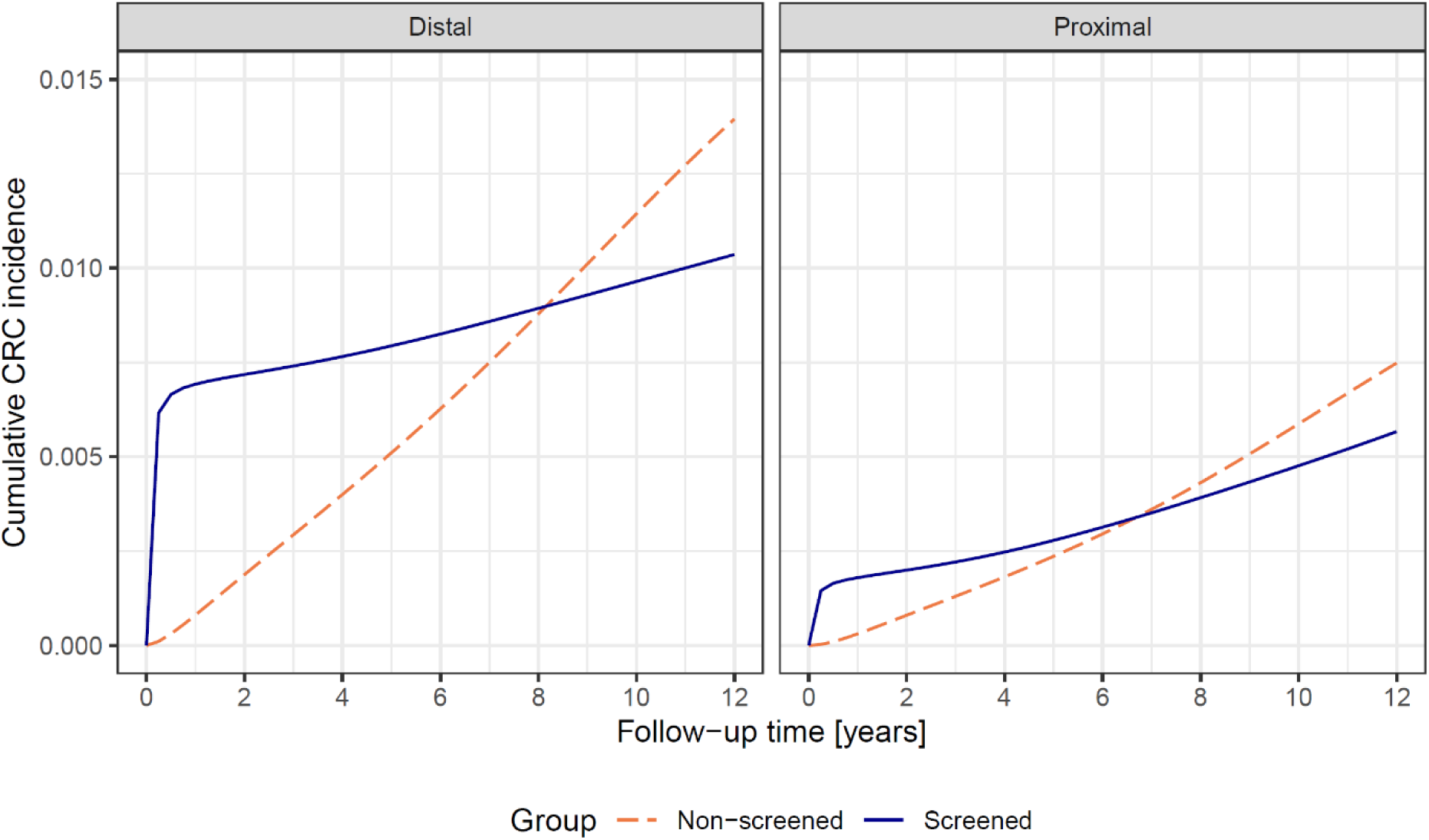
Results of the per-protocol analysis for the cohort design with alignment at time zero: adjusted cumulative CRC incidence in the screening and the control arm

## Supplement 3: Cohort and case-control study without alignment at time zero for different baseline years

As mentioned in the methods section, for the study designs without alignment at time zero, we selected individuals from the source population entering the cohort in 2009. In sensitivity analyses, we varied the baseline year, i.e. individuals entering the cohort in 2010 and 2011, respectively. The respective results are shown in Table S1 and Figure S1 for the cohort study and in Table S2 for the case-control study. For comparison, also the results of the base case analysis (baseline year 2009) are shown.

**Table S1:** Results of cohort study designs without alignment at time zero for different baseline years.

| Baseline year |  | Control group | Screening group | Adjusted relative risk | (95% CI) |
| --- | --- | --- | --- | --- | --- |
|  | Number at risk | 200,000 | 200,000 |  |  |
|  | Number of CRC cases |  |  |  |  |
| 2009 | Distal, n | 2,472 | 829 | 0.35 | (0.32-0.39) |
|  | Proximal, n | 1,290 | 823 | 0.63 | (0.57-0.69) |
| 2010 | Distal, n | 2,101 | 709 | 0.35 | (0.32-0.39) |
|  | Proximal, n | 1,131 | 702 | 0.61 | (0.56-0.68) |
| 2011 | Distal, n | 2,048 | 642 | 0.33 | (0.30-0.37) |
|  | Proximal, n | 1,056 | 640 | 0.54 | (0.49-0.59) |

**Figure S1:**
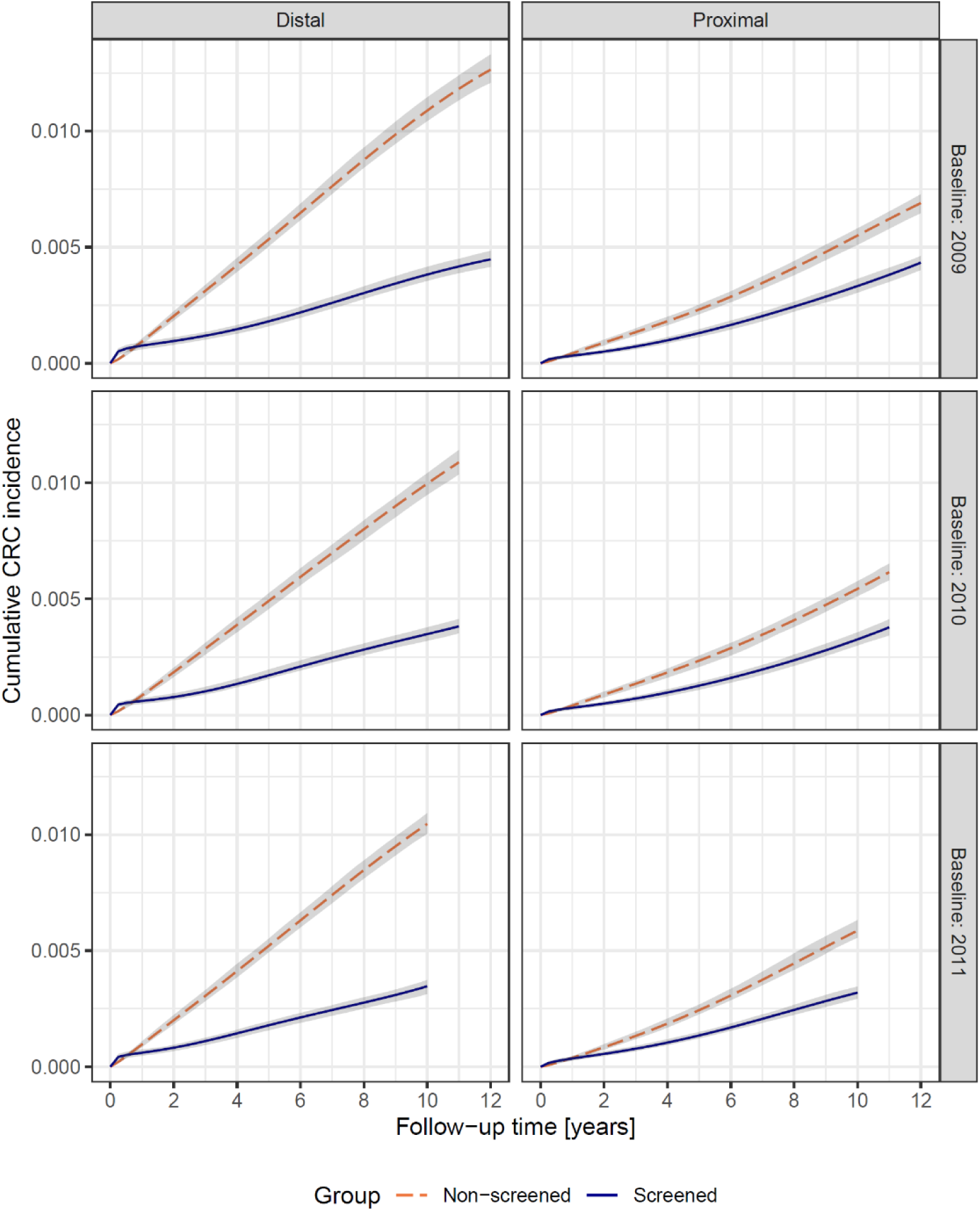
Adjusted cumulative incidence functions for distal and proximal CRC from the cohort study design without alignment at time zero for different baseline years.

**Table S2:**
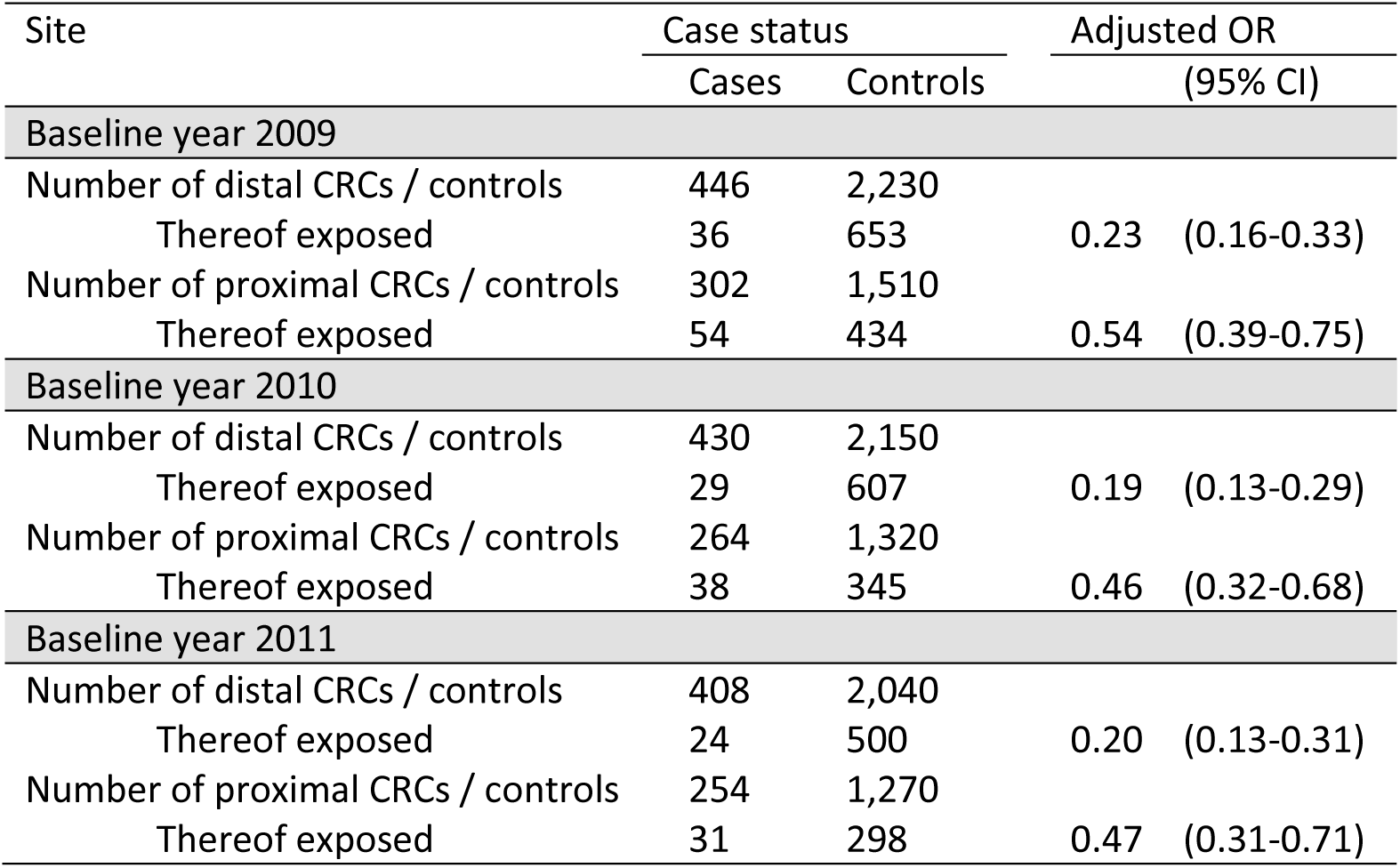
Results of case-control designs without alignment at time zero for different baseline years.

## Supplement 4: Cohort and case-control study without alignment at time zero: considering both screening and diagnostic colonoscopy for the assignment to exposure groups

As mentioned in the methods section regarding the study designs without alignment at time zero, only screening colonoscopies were considered for the assignment to exposure groups in the base case analysis. In a sensitivity analysis, we considered both screening and diagnostic colonoscopies for the exposure assignment. The respective results are shown in Table S4 and Figure S2 for the cohort study design and in Table S5 for the case-control study.

**Table S4:**
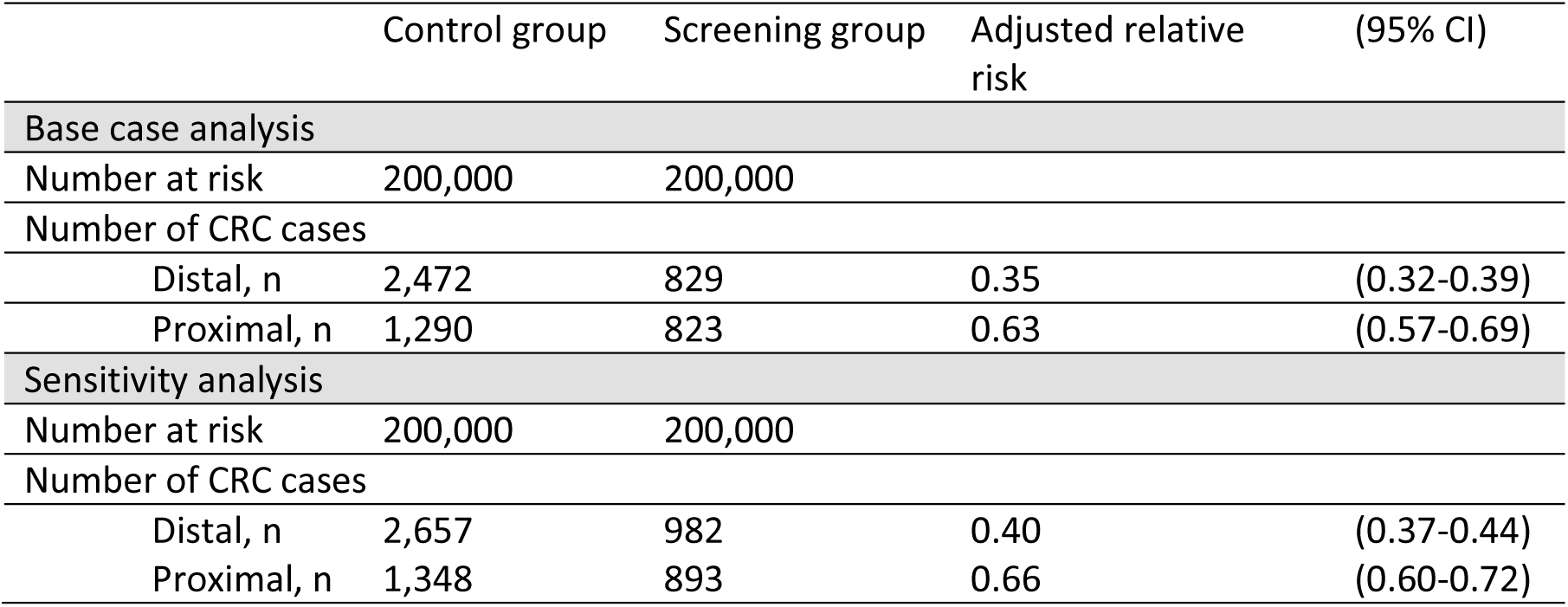
Results of cohort study designs without alignment at time zero: sensitivity analyses considering both screening and diagnostic colonoscopy for the assignment to exposure groups. For comparison, also the results of the base case analysis are shown.

|  | Control group | Screening group | Adjusted relative risk | (95% CI) |
| --- | --- | --- | --- | --- |
| **Base case analysis** |  |  |  |  |
| Number at risk | 200,000 | 200,000 |  |  |
| Number of CRC cases |  |  |  |  |
| Distal, n | 2,472 | 829 | 0.35 | (0.32-0.39) |
| Proximal, n | 1,290 | 823 | 0.63 | (0.57-0.69) |
| **Sensitivity analysis** |  |  |  |  |
| Number at risk | 200,000 | 200,000 |  |  |
| Number of CRC cases |  |  |  |  |
| Distal, n | 2,657 | 982 | 0.40 | (0.37-0.44) |
| Proximal, n | 1,348 | 893 | 0.66 | (0.60-0.72) |

**Figure S2:**
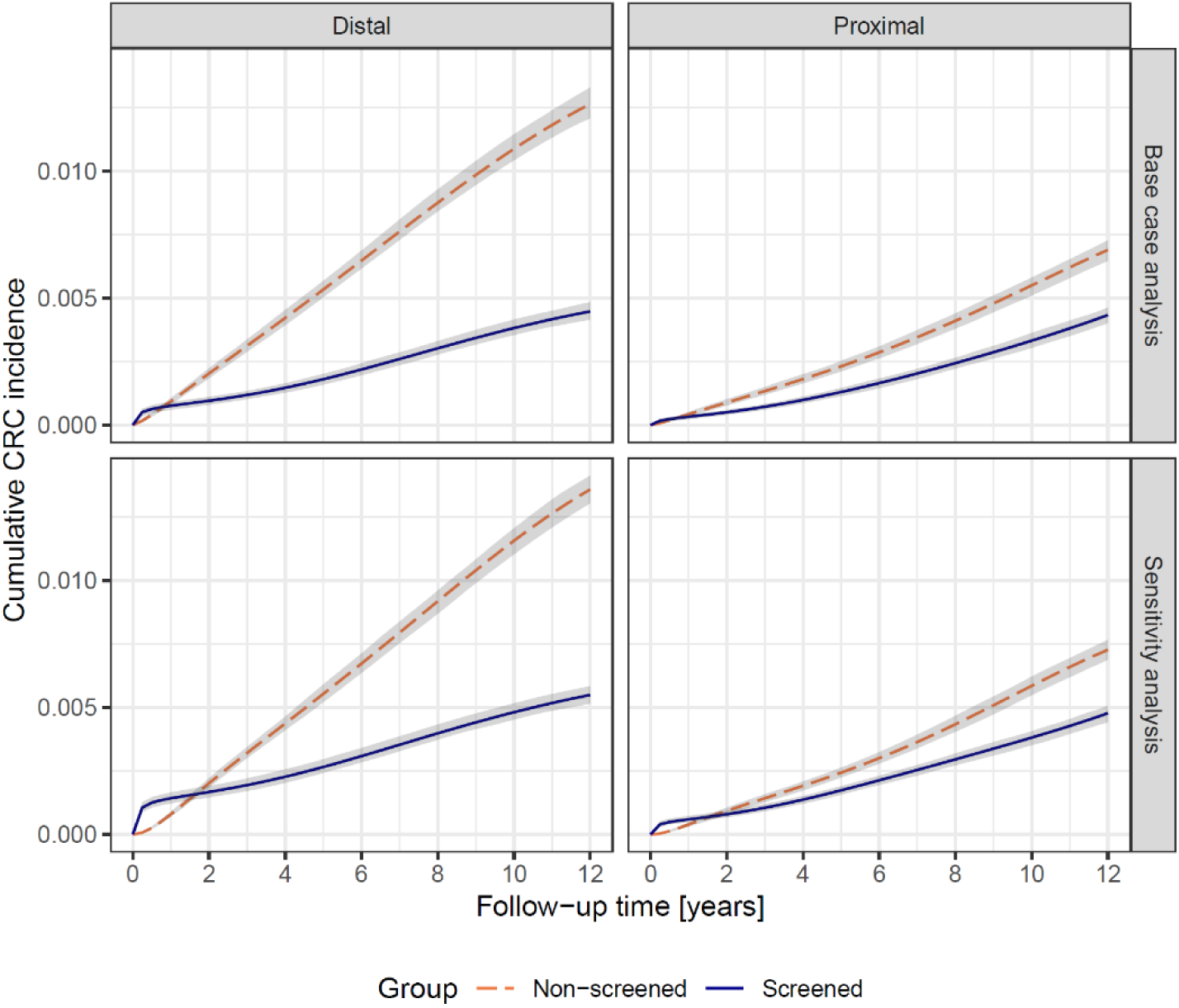
Adjusted cumulative incidence functions for distal and proximal CRC from the cohort study design without alignment at time zero: sensitivity analysis considering both screening and diagnostic colonoscopy for the assignment to exposure groups. For comparison, also the cumulative incidence functions of the base case analysis are shown.

**Table S5:**
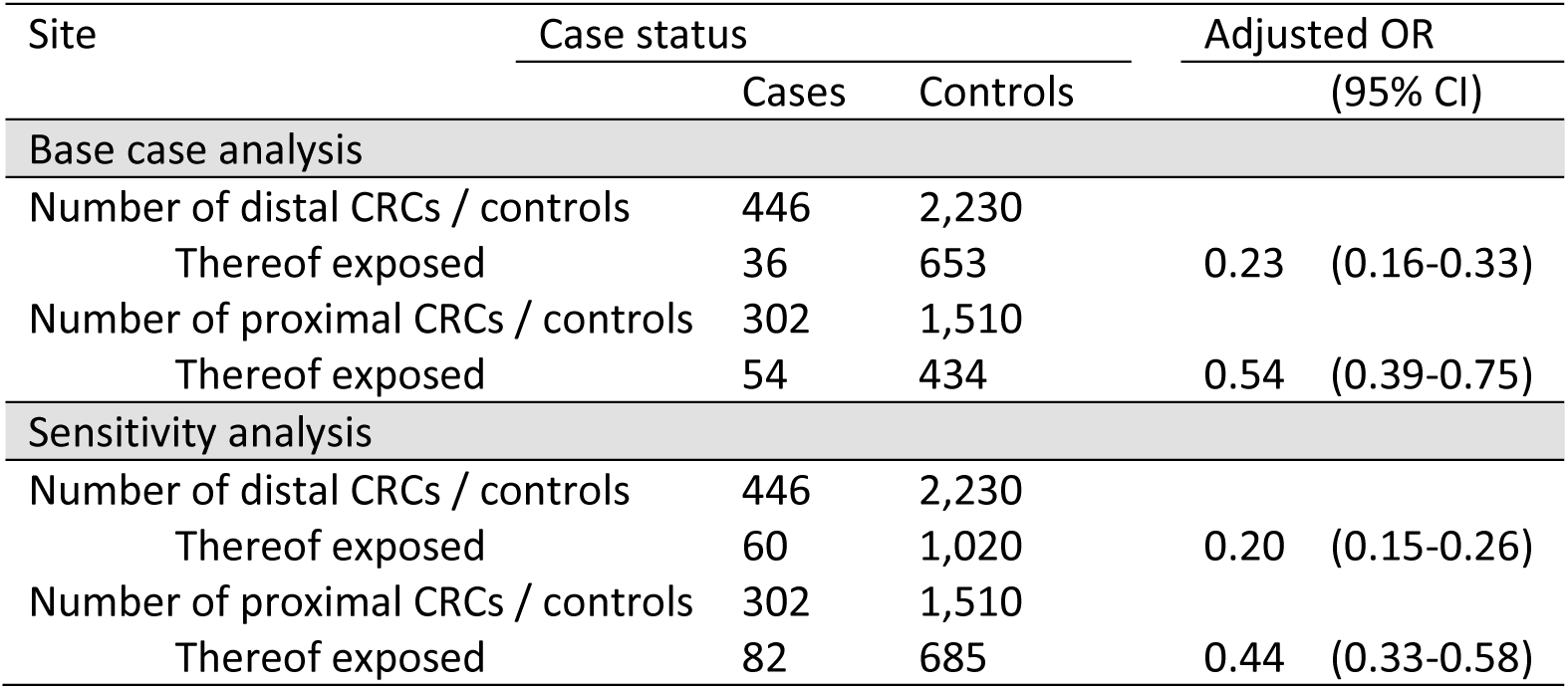
Results of case-control designs without alignment at time zero: sensitivity analyses considering both screening and diagnostic colonoscopy for the assignment to exposure groups. For comparison, also the results of the base case analysis are shown.

## Supplement 5: Post-colonoscopy CRC diagnoses

For the cohort analysis with alignment at time zero, we quantified the occurrence of post-colonoscopy CRC diagnoses occurring in the screening arm and assessed their site distribution. CRC diagnoses with a screening colonoscopy in the same calendar quarter or in the 180 days before CRC diagnosis were considered screen-detected and were not counted as post-colonoscopy CRC. The frequencies and percentages are given in the below Table:

| Site | N | % |
| --- | --- | --- |
| Distal CRC | 541 | 39.3 |
| Proximal CRC | 633 | 46.0 |
| Both/unknown | 203 | 14.7 |
| Total | 1377 |  |

## Supplement 6: Table of baseline covariates by study design and screening group

| Baseline covariate | Cohort designs |  |  |  | Case-control designs |  |  |  |
| --- | --- | --- | --- | --- | --- | --- | --- | --- |
|  | With alignment |  | Without alignment |  | With alignment |  | Without alignment |  |
|  | Screened<br>n (%) | Not screened<br>n (%) | Screened<br>n (%) | Not screened<br>n (%) | Screened<br>n (%) | Not screened<br>n (%) | Screened<br>n (%) | Not screened<br>n (%) |
| Age |  |  |  |  |  |  |  |  |
| Median (Q1-Q3) | 61 (57-65) | 62 (58-66) | 63 (59-67) | 62 (58-66) | 63 (59-67) | 64 (60-67) | 62 (58-66) | 63 (59-67) |
| Mean (SD) | 61.1 (4.53) | 61.7 (4.48) | 62.9 (4.14) | 61.9 (4.57) | 62.7 (4.41) | 63.1 (4.29) | 61.9 (4.48) | 62.8 (4.39) |
| Female | 95,401 (49.7) | 656,855 (52.9) | 114,840 (57.4) | 112,233 (56.1) | 9,617 (39.0) | 73,532 (44.3) | 1,367 (41.1) | 4,747 (45.7) |
| Education |  |  |  |  |  |  |  |  |
| No degree/unknown | 104,108 (54.2) | 755,919 (60.9) | 127,838 (63.9) | 124,114 (62.1) | 15,542 (63.1) | 113,015 (68.1) | 1,820 (54.7) | 6,609 (63.6) |
| Basic or secondary degree | 47,508 (24.7) | 293,925 (23.7) | 41,368 (20.7) | 46,164 (23.1) | 4,775 (19.4) | 31,134 (18.8) | 741 (22.3) | 2,223 (21.4) |
| Higher education | 40,438 (21.1) | 191,227 (15.4) | 30,794 (15.4) | 29,722 (14.9) | 4,332 (17.6) | 21,780 (13.1) | 769 (23.1) | 1,566 (15.1) |
| Codes indicating obesity | 38,928 (20.3) | 254,280 (20.5) | 41,836 (20.9) | 42,397 (21.2) | 4,884 (19.8) | 33,796 (20.4) | 656 (19.7) | 2,259 (21.7) |
| Codes for alcohol abuse | 5,257 (2.7) | 51,255 (4.1) | 4,992 (2.5) | 8,352 (4.2) | 647 (2.6) | 6,240 (3.8) | 94 (2.8) | 461 (4.4) |
| Codes for heavy smoking | 12,140 (6.3) | 99,370 (8.0) | 10,276 (5.1) | 15,686 (7.8) | 1,384 (5.6) | 11,799 (7.1) | 225 (6.8) | 829 (8.0) |
| At least one other preventive service before baseline | 172,324 (89.7) | 904,168 (72.9) | 192,416 (96.2) | 159,476 (79.7) | 21,763 (88.3) | 119,433 (72.0) | 2,906 (87.3) | 7,611 (73.2) |
| Diabetes |  |  |  |  |  |  |  |  |
| Drug-treated diabetes | 13,453 (7.0) | 114,282 (9.2) | 12,986 (6.5) | 17,831 (8.9) | 1,930 (7.8) | 16,313 (9.8) | 249 (7.5) | 980 (9.4) |
| Diabetes with end organ damage | 597 (0.3) | 7,643 (0.6) | 589 (0.3) | 1,322 (0.7) | 89 (0.4) | 952 (0.6) | 9 (0.3) | 52 (0.5) |
| Codes for family history of CRC | 423 (0.2) | 1,077 (0.1) | 851 (0.4) | 236 (0.1) | 41 (0.2) | 113 (0.1) | 5 (0.2) | 8 (0.1) |
| Menopausal hormone therapy<sup>§</sup> (percent refer to women) | 37,393 (39.1) | 158,937 (24.2) | 52,466 (45.6) | 34,595 (30.8) | 3,668 (38.1) | 17,537 (23.8) | 479 (35.0) | 909 (19.1) |
| Antiplatelet therapy |  |  |  |  |  |  |  |  |
| Only acetylsalicylic acid | 7,850 (4.1) | 58,917 (4.7) | 8,697 (4.3) | 9,646 (4.8) | 1,167 (4.7) | 8,878 (5.4) | 143 (4.3) | 468 (4.5) |
| Dual | 1,413 (0.7) | 11,028 (0.9) | 1,392 (0.7) | 1,749 (0.9) | 228 (0.9) | 1,601 (1.0) | 24 (0.7) | 109 (1.0) |
| Without acetylsalicylic acid | 2,977 (1.6) | 24,219 (2.0) | 3,319 (1.7) | 3,965 (2.0) | 446 (1.8) | 3,473 (2.1) | 62 (1.9) | 224 (2.2) |

<sup>§</sup>: We used a sensitive definition of MHT (at least one prescription in the three years before baseline) which explains the relatively high proportion of women using MHT.

## Supplement 7: Structural explanation of the bias inherent to study designs using pre-baseline information for the assignment to exposure groups

For simplicity, we divide time into three periods 𝑡 ∈ {−1, 0, 1} with 𝑡 = −1 being the pre-baseline period, 𝑡 = 0 the baseline and 𝑡 = 1 the post-baseline or follow-up period. Let 𝐸_𝑡_ ∈ {0, 1} described a person’s exposure to screening colonoscopy at time 𝑡. Let 𝑃_𝑡_ ∈ {0, 1} indicate the presence of colorectal precursors at time 𝑡 and 𝐶_𝑡_ ∈ {0, 1} the onset of preclinical CRC by time 𝑡. Let 𝑌_𝑡_ ∈ {0, 1} indicate a diagnosis of colorectal cancer by time 𝑡. The box around 𝑌_−1_ indicates that inclusion in the analysis population is based on previous CRC diagnoses, i.e. individuals are only included in the study population if 𝑌_−1_ = 0.

**Figure S3:**
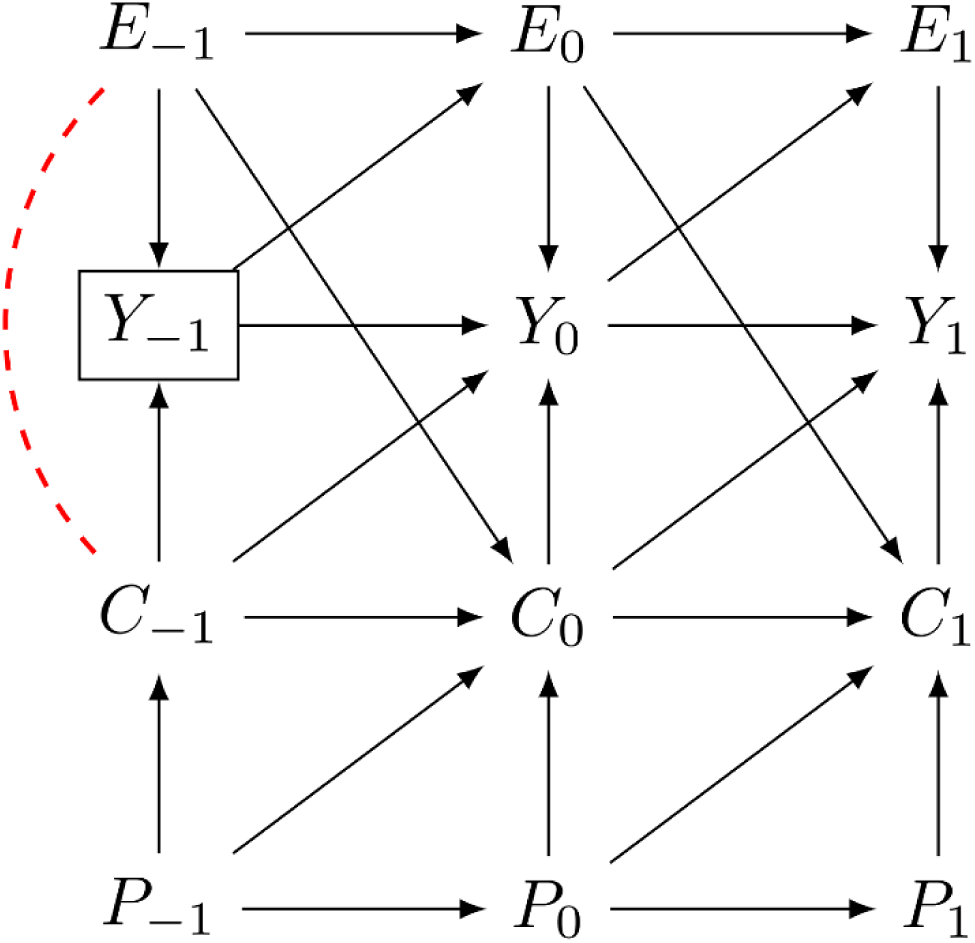
DAG of bias resulting from violation of alignment at time zero in the form of exposure assessment based on pre-baseline information.

As shown in Figure S3, at time point 𝑡 the causal mechanism that leads to a diagnosis of CRC is as follows: Precursors 𝑃_𝑡_ lead to the development of CRC 𝐶_𝑡_, which in turn progress to the outcome of interest, CRC diagnosis 𝑌_𝑡_. At the same time, exposure to screening colonoscopy 𝐸_𝑡_ leads to CRC diagnosis 𝑌_𝑡_ at the same time point, if the disease is present. Furthermore, exposure at time 𝑡 prevents disease onset at the later time 𝑡 + 1 by removing precursor stages present at time 𝑡.

Importantly, the variable 𝑌_𝑡_ is a collider variable on the path 𝑃_𝑡_ → 𝐶_𝑡_ → 𝑌_𝑡_ ← 𝐸_𝑡_. When cohort selection is based on this collider, a non-causal association is introduced between 𝐸_𝑡_ and 𝐶_𝑡_. If the cohort selection process excludes individuals with CRC diagnosis before baseline (𝑌_−1_) while including individuals with past exposure 𝐸_−1_ in the exposed group of the analysis dataset, the unexposed group appears to have a higher CRC incidence. Individuals who were screened in the past and had prevalent CRC received a diagnosis and were filtered out of the study cohort. Individuals who were screened in the past and did not have prevalent CRC are included in the exposed group. No such selection takes place in the unexposed group, where individuals must not have had any screening colonoscopy before baseline. Therefore, there is a non-causal association between exposure before baseline and prevalent, undiagnosed CRC before baseline. This non-causal association means that there are now open non-causal paths from exposure before baseline to the study outcome at later time points. The resulting bias, therefore, can be expressed as a form of collider stratification bias. The analysis with alignment at time zero would also exclude individuals with previous exposure 𝐸_−1_ = 1 and would aim to estimate the effect of exposure at time zero (𝐸_0_) on subsequent outcomes. The analysis without alignment at time zero on the other hand does not exclude individuals with previous exposure and instead aims to estimate the effect of the composite exposure 𝐸_misaligned_ = {𝐸_−1_ = 1 or 𝐸_0_ = 1}, which does not correspond to a causal question since past exposure cannot be intervened upon.

The strength of the bias will depend on the prevalence of 𝐶_−1_. If, conceptually, the prevalence of CRC before baseline were to approach zero, no such selection would take place. In the age group under study here, the prevalence of proximal CRC before baseline will be much lower than the prevalence of distal CRC before baseline, which means that this bias will impact the effect estimate for distal CRC more severely.

## Supplement 8: Illustration of the mechanism underlying the misallocation of screen-detected CRCs in case-control studies without alignment at time zero

Figure S4 illustrates the mechanism that underlies the misallocation of screen-detected CRCs in case-control studies without alignment at time zero, resulting in an overestimate of the effectiveness of screening colonoscopy. First, let us imagine a hypothetical RCT investigating the effectiveness of screening colonoscopy on CRC incidence. At baseline, screening-na1ve persons are randomly assigned to either the screening or the control arm. Analysing this data as a case-control study without alignment at time zero would mean that for CRCs occurring in both arms, it is assessed whether they had a colonoscopy *before* CRC diagnosis. Given that screen-detected CRCs did not have a colonoscopy *before* CRC diagnosis, they are assigned (post-baseline, i.e. after randomization) to the control arm and are thereby classified as unexposed. This overestimates the effectiveness of screening given that CRCs accumulate in the control group (unexposed group). Given that screen-detected CRCs are more frequent in the distal colorectum, the resulting bias affects distal CRC more severely than proximal CRC.

**Figure S4:**
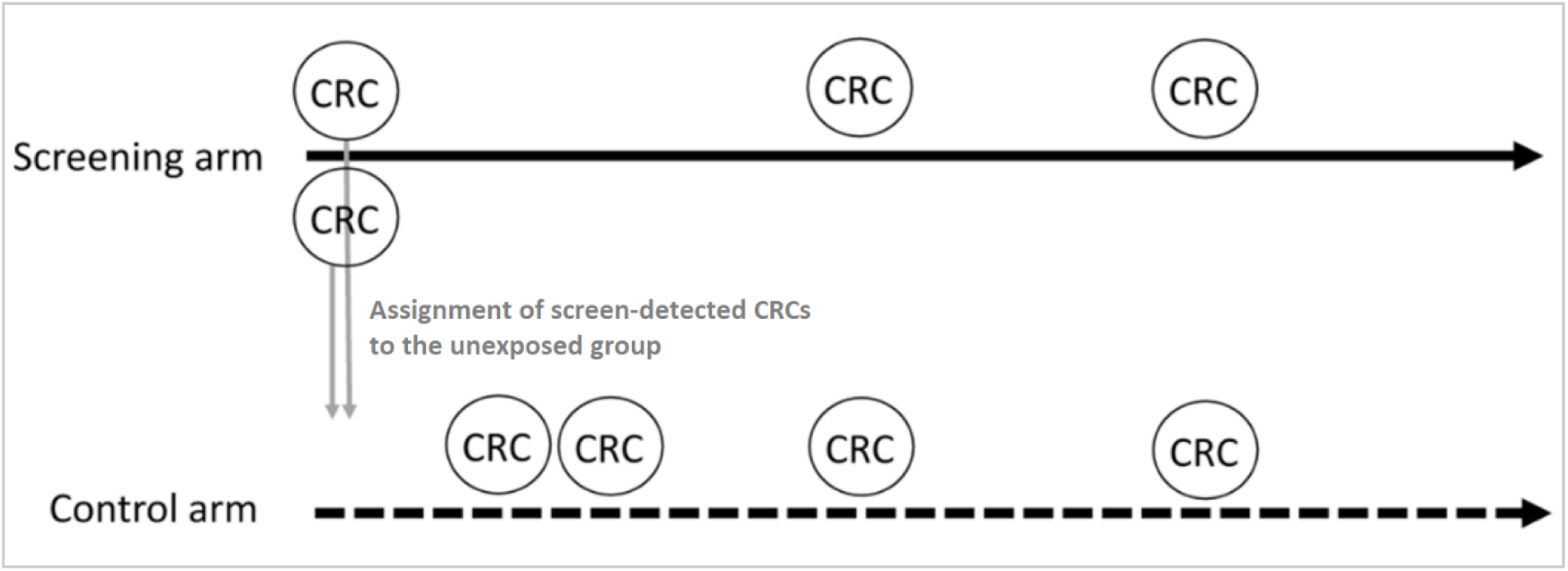
Illustration of the mechanism of misallocation of screen-detected CRCs in case-control studies without alignment at time zero

Of note, in published case-control studies investigating the effectiveness of screening colonoscopy based on primary data, selection bias in the control arm (higher prevalence of screening colonoscopy as compared to the general population) can—as an additional mechanism—also contribute to overestimating the effectiveness of screening colonoscopy, but it is not expected that this bias leads to a difference in the effectiveness by site.

In our case-control study without alignment at time zero, there was a second mechanism leading to overestimating the effectiveness of screening colonoscopy due a compromise we had to make because of the left truncation of our data. Specifically, we had to select CRC cases diagnosed in 2018-2020 from those entering the cohort in 2009 (see methods section) in order to be able to assess exposure in the 10 years prior to CRC diagnosis. CRCs diagnosed between 2009 and 2017 in the context of screening, which are more often in the distal than in the proximal colon, were not included in the final set of cases, i.e. distal CRCs exposed to screening colonoscopy were underrepresented in the final set of cases. We conducted additional analyses to disentangle the effect of both mechanism (data not shown), which did not change our conclusion, i.e. that the mechanism described in Figure S4 (also) leads to an artificial difference in the effectiveness of colonoscopy by site.

Further sources of bias due to misalignment in case-control designs may exist, e.g. when eligibility and covariates are assessed at time zero but exposure assessment uses information from after time zero. For example, exposure after time zero may be influenced by time-dependent confounders that changed since baseline.

## Supplement 9: Bias due to post-baseline information for exposure assignment in a cohort study

In the cohort study by Nishihara et al. the assessment of eligibility criteria (e.g. no prior cancer except for nonmelanoma skin cancer, no prior endoscopy) as well as the start of follow-up was in 1988 (baseline) ^2^. As part of a questionnaire administered every 2 years, participants were then asked whether they had undergone either sigmoidoscopy or colonoscopy and, if so, the reason for the investigation and whether there was a diagnosis of colorectal polyps. This means that the assignment to exposure groups used information after the assessment of eligibility and the start of follow-up, and it was updated every two years, i.e. post-baseline information was used to determine exposure. The outcome was the incidence of colorectal cancer, which was compared between participants without a lower endoscopy (control group), participants with a polypectomy, participants with a negative sigmoidoscopy and participants with a negative colonoscopy.

The mechanism described for the case-control study without alignment at time zero also applies to this design. In each two-year time interval CRCs detected in persons who had their first colonoscopy during this two-year time interval are—per definition—assigned to the unexposed group as they had no colonoscopy prior to CRC diagnosis. This overestimates the effectiveness of screening because CRCs are filtered to the unexposed group. As the majority of screen-detected CRCs are in the distal colon, this bias predominantly affects CRCs in the distal colon and thus leads to an artificial difference in the effectiveness of colonoscopy by site.

## Notes

### Competing Interest Statement

The authors have declared no competing interest.

### Funding Statement

BIPS intramural funding

### Author Declarations

In Germany, the utilisation of health insurance data for scientific research is regulated by the Code of Social Law. All involved health insurance providers as well as the German Federal Office for Social Security and the Senator for Health, Women and Consumer Protection in Bremen as their responsible authorities approved the use of GePaRD data for this study. Informed consent for studies based on claims data is required by law unless obtaining consent appears unacceptable and would bias results, which was the case in this study. According to the Ethics Committee of the University of Bremen studies based on GePaRD are exempt from institutional review board review.

### Summary of Updates

This version has been updated to incorporate reviewer comments from the second round of blinded peer-review (minor revision).

